# The early dynamics of the SARS-CoV-2 epidemic in Portugal

**DOI:** 10.1101/2021.02.22.21252216

**Authors:** V Borges, J Isidro, NS Trovão, S Duarte, H Cortes-Martins, H Martiniano, I Gordo, R Leite, L Vieira, Portuguese network for SARS-CoV-2 genomics (Consortium), R Guiomar, JP Gomes

## Abstract

**Background:** Genomic surveillance of SARS-CoV-2 in Portugal was rapidly implemented by the National Institute of Health in the early stages of the COVID-19 epidemic, in collaboration with more than 50 laboratories distributed nationwide. This unprecedented collaborative effort culminated in the generation of 1275 SARS-CoV-2 genome sequences, which represent 15.5% of all confirmed cases in March 2020, making Portugal one of the countries generating the highest volumes of SARS-CoV-2 genomic data during early COVID-19 pandemic.

**Methods:** We reconstructed and characterized the spatio-temporal dynamics of SARS-CoV-2 introductions and early dissemination in Portugal using recent phylodynamic models that allow integration of individual-based travel history, in order to obtain a more realistic reconstruction of the viral dynamics.

**Results:** We detected at least 277 independent SARS-CoV-2 introductions, mostly from European countries (namely the United Kingdom, Spain, France, Italy and Switzerland), which was broadly consistent with the available travel history data, as well as with the countries with most frequent connectivity and/or with the highest number of Portuguese immigrants. Although most introductions were estimated to have occurred during the last week of February and the first week of March 2020, it is likely that SARS-CoV-2 was silently circulating in Portugal several weeks before the first confirmed local cases on March 2, 2020.

**Discussion and Conclusion:** While the implemented preventive and early control measures seem to have been successful in mitigating community transmission from most independent introductions, our results suggest that their earlier implementation could have largely minimized the number of introductions and subsequent virus expansion. Here we lay the foundation for genomic epidemiology of SARS-CoV-2 in Portugal, and highlight the need for systematic, continuous and geographically-representative genomic surveillance to guide national and international public health authorities toward the characterization and control of SARS-CoV-2 circulating diversity.

## INTRODUCTION

SARS-CoV-2 (Severe Acute Respiratory Syndrome Coronavirus 2), the causative agent of COVID-19, is a novel betacoronavirus that was first reported in December 2019 in Wuhan, China (**Zhou et al, 2020; Wu et al, 2020**). It was declared a global public health emergency that, as of February 17, 2020, has caused more than 109 million cases and two million deaths worldwide (**WHO, 2021; Dong et al, 2020**). In order to control the virus arrival and spread, many countries adopted rigid public health measures, including complete border closures and general lockdowns, with tremendous consequences at economic and social levels. At the early stages of an epidemic, the success of public health measures is particularly dependent on their timely implementation, which requires comprehensive diagnosis/surveillance systems that are able to efficiently trace where the virus is being introduced and circulating (**Alm et al, 2020; Nadeau et al, 2020; Gámbaro et al, 2020; du Plessis et al, 2021**). Taking advantage of the recent extraordinary advances in sequencing technologies, modern surveillance systems are progressively relying on genomic epidemiology as a crucial tool for outbreak investigation and for tracking virus evolution and spread (**Fauver et al, 2020; Lemieux et al, 2020; du Plessis et al, 2021**). Genomic surveillance of SARS-CoV-2 can be particularly useful for: i) understanding the contribution of “new introductions” versus “local transmission” to the number of new cases at continent/country/regional levels; ii) evaluating the impact of non-pharmaceutical interventions on the outcomes of transmission chains; iii) characterizing the genetic variability that may negatively affect molecular diagnostic tests; iv) identifying and monitoring genetic variability affecting antigens and targets of antiviral drugs with potential impact on the development/effectiveness of prophylactic (vaccines) and therapeutic measures; v) investigating potential associations between genetic variants and infectious load, patient immunological status, clinical outcomes (e.g., infection duration, disease severity, etc) (**Alm et al, 2020**). Acting as the National Reference Laboratory for SARS-CoV-2, the Portuguese National Institute of Health (INSA) Doutor Ricardo Jorge rapidly established a genome-based molecular surveillance strategy for SARS-CoV-2 in Portugal, setting up a large nationwide network involving more than 50 laboratories. A website (https://insaflu.insa.pt/covid19) was launched, providing updated data regarding the analysis of the SARS-CoV-2 genetic diversity and geotemporal dynamics. Also, “situation reports” with major highlights are being released periodically to participating laboratories, national and regional public health authorities and other stakeholders. Despite all the advantages of genomic surveillance, the uneven geographic sampling of viral genomes can severely skew phylogeographic inferences based on discrete trait ancestral reconstruction (**Lemey et al, 2020**), therefore hindering the ability to accurately trace the seeding and dissemination patterns of SARS-CoV-2. The COVID-19 pandemic has been characterized by an unprecedented amount of genomic data and associated metadata, such as information on the patients’ recent movements prior to having developed any symptoms.

In the present study, we reconstruct and characterize the spatio-temporal dynamics of SARS-CoV-2 introductions and early dissemination in Portugal using newly developed phylodynamic models that allow integration of individual-based travel history, in order to obtain a more realistic reconstruction of the viral dynamics (**Lemey et al, 2020**). This includes inferences of the timeliness of the very first introductions, geographic location of ancestral lineages and the contribution of detected introductions on the epidemic evolution.

## MATERIAL AND METHODS

### Sample characterization

Samples used in this study were collected as part of the ongoing national SARS-CoV-2 laboratory surveillance conducted by the National Institute of Health (INSA) Doutor Ricardo Jorge, Portugal. SARS-CoV-2 positive samples (either clinical specimens or RNA) were provided by a nationwide network, consisting of more than 50 laboratories, that was established at the beginning of the epidemic in Portugal. Anonymized date of sample collection, date of illness onset and travel history were provided by laboratories and Regional and National Health Authorities. Geographical data presented in this study refers to the Region (“Health Administration region”) the patients’ residence or, when no information is available (for a small proportion of cases), to the Region of exposure or of the hospital/laboratory that collected/sent the sample.

### SARS-CoV-2 amplicon-based genome amplification and sequencing

SARS-CoV-2 positive RNA samples were subjected to genome sequencing using a whole-genome amplification strategy with tiled, multiplexed primers (**Quick et al, 2017**) and the ARTIC Consortium protocol (https://artic.network/ncov-2019; https://www.protocols.io/view/ncov-2019-sequencing-protocol-bbmuik6w), with slight modifications. In brief, after cDNA synthesis using SuperScript™ IV First-Strand Synthesis System kit (Invitrogen™, catalog: 18091050) with random hexamers from 11 μL of RNA (exactly as described in ARTIC Network protocol), targeted amplification was performed with 2·5 μL of cDNA using NEBNExt® Q5® HotStart HiFi Master Mix (12·5 μL per reaction) (New England BioLabs, catalog: M0544S) with two pools of tiling primers (A and B) separately. Primers versions V1 and V2 (aliquots kindly provided by ARTIC Network team) were used for the first 243 samples of this study, while the V3 primers (with a total of 218 primers) were applied to all samples afterwards (all versions available here: (https://github.com/artic-network/artic-ncov2019/tree/master/primer_schemes/nCoV-2019). The final concentration per primer (V3 version) was ∼0·013µM (1·4 µM per pool) in a 25 μL total reaction volume. PCR amplification parameters were: 30s at 98°C, 35 cycles of 15s at 98°C and 5 min at 65°C (for the first 762 samples) or 63°C (afterwards), and final extension for 5 min at 65/63°C. Amplicons were visualized on a 1% agarose gel, tubes A and B were pooled per sample, and subjected to clean up with Agencourt AMPure XP (Beckman Coulter, catalog: A63880) using a 1:1 volume ratio. Purified amplicons were quantified using Qubit fluorometer (Thermo Fisher Scientific) and normalized to a concentration of 0·4 ng/ul. Dual-indexed sequencing libraries were prepared using Illumina Nextera XT DNA Library Prep Kit (Illumina). Pooling, denaturation and dilution of bead-based normalized libraries was performed according to the manufacturer’s instructions for the MiSeq or NextSeq 550 systems (Illumina). A 1% spike-in PhiX genome library (Illumina) was used as internal quality control. Libraries were sequenced using 250 base pairs (bp) (MiSeq) or 150bp (NextSeq 550) paired-end reads targeting ∼1M reads per sample.

### Genome assembly and sequence curation

Analysis of sequence read data was conducted using the bioinformatics pipeline implemented in INSaFLU (https://insaflu.insa.pt/; https://github.com/INSaFLU), which is a web-based (and also locally installable) platform for amplicon-based next-generation sequencing data analysis (**Borges et al, 2018**). Briefly, the core bioinformatics steps (documented in **Borges et al, 2018** and https://insaflu.readthedocs.io/) involved: i) raw NGS reads quality analysis and improvement using FastQC; (https://www.bioinformatics.babraham.ac.uk/projects/fastqc) and Trimmomatic (http://www.usadellab.org/cms/index.php?page=trimmomatic), respectively (read’s ends were cropped 30bp for primer clipping); ii) draft *de novo* assembly using SPAdes (http://cab.spbu.ru/software/spades/) followed by classification and contigs assignment of Human Betacoronavirus; and, iii) reference-based mapping, consensus generation and mutation detection using the multisoftware tool Snippy (https://github.com/tseemann/snippy), using the Wuhan-Hu-1/2019 genome sequence (https://www.ncbi.nlm.nih.gov/nuccore/MN908947) as reference. The first 40bp and end 100bp were discarded and consensus sequences were exclusively included in the study when >70% of the genome was covered by at least 10-fold. For samples with coverage drop below 10-fold, fine-tuned consensus sequence curation was performed as follows: i) undefined bases (“N”) were placed in genome regions with depth of coverage below 10 using a python script (https://github.com/rfm-targa/BioinfUtils/blob/master/msa_masker.py), and “N” regions at the sequence ends were trimmed to avoid releasing sequences starting or ending with “N”;ii) all regions with depth coverage below 10 were visually inspected in the Integrative Genomics Viewer (http://www.broadinstitute.org/igv) available *in situ* at INSaFLU, and when mutations were detected and validated within these low coverage regions, they were inserted in the consensus sequence to avoid disrupting/biasing the phylogenetic signal; iii) when a mutation was validated within a “N” region with <=100bp, the whole region was inserted (as long as the region was covered by at least one read); when a mutation was validated within a “N” region with >100bp, the mutation was inserted together with additional 20bp (10bp from each flanking side) to improve the downstream sequence alignment.

### Classification by clades and lineages

We explored the diversity of INSA sequences using a variety of nomenclature strategies, namely Nextstrain (using https://clades.nextstrain.org/; 9 November 2020), GISAID (https://www.gisaid.org/; 23 July 2020) and Phylogenetic Assignment of Named Global Outbreak LINeages (cov-lineages.org) (https://pangolin.cog-uk.io/; 16 October 2020) (**Rambaut et al, 2020**). While Nextstrain and GISAID clade nomenclatures provide a less detailed categorisation of globally circulating diversity, cov-lineages.org classification is focused on identifying highly specific lineages that are actively transmitting in the population (**Rambaut et al, 2020**). Classification is provided in Supplementary material (**Table S1**).

### Assessment of genome sampling by country

To assess the contribution of each country to the set of publicly available SARS-CoV-2 genomes and to determine the proportion of the number of genomes on the total number of reported COVID-19 cases (genome sampling) of a given country during the study period (until March 31, 2020), we obtained the number of cases per country from the COVID-19 Data Repository by the Center for Systems Science and Engineering (CSSE) at Johns Hopkins University (https://github.com/CSSEGISandData/COVID-19/blob/master/csse_covid_19_data/csse_covid_19_time_series/time_series_covid19_confirmed_global.csv) and the number of genomes from GISAID (by August 8, 2020). Only the genomes with collection date until March 31, 2020 were considered. When a given genome only indicated the month of collection, but not the day, it was added to the sum of the last day of the respective month. When the number of genomes on a given day was higher than the number of cases, the number of cases was considered for graphical representation. Final data on the assessment of genome sampling by country is provided in Supplementary material (**Table S2**).

### Selecting a genomic background dataset

For the phylogenetic analyses, we downloaded full-length viral genome sequences from GISAID (https://www.gisaid.org/) on August 6, 2020 with collection dates before April 1, 2020 (**Table S2**). For computational efficiency in the downstream operations, we analysed the A and B lineages seperately. Multiple sequence alignments with a reference genome (MN908947.3) were performed using MAFFT v7.458 with parameter --addfragments (Katoh et al, 2009). Sequences with fewer than 75% unambiguous bases were excluded, as well as duplicate sequences defined as having identical nucleotide composition, collected on the same date and in the same country. The resulting dataset was trimmed at the 5’ and 3’ ends resulting in a multi-sequence alignment with 29780 nucleotides. Sequences with date information at the year-level were also excluded. This dataset was subjected to multiple iterations of phylogeny reconstruction using IQ-TREE multicore software version 1.6.12 (**Nguyen et al, 2015**) with parameters -m GTR+G, and exclusion of outlier sequences whose genetic divergence and sampling date were incongruent using TempEst software version 1.5.2 (**Rambaut et al, 2016**), resulting in 1632 and 22124 sequences for the A and B datasets, respectively. GISAID acknowledgment table for the background dataset is provided as Supplementary Material (**Table S3**).

### Subsampling strategy

The magnitude of the B datasets prohibits a full Bayesian inference approach in a reasonable timeframe. To overcome this constraint, we used a subsampling strategy that removes sequences such that monophyletic clusters that consist entirely of sequences from a particular country are represented by a single sequence. The excess sequences in a country-specific monophyletic clade do not contribute any additional information to the between-country diffusion process we aim to infer (**Hong et al, 2020**). This process resulted in a dataset with 13489 sequences (B_CS). Despite the almost 40% downsampling in the B lineage dataset, its size is still excessive for timely computational inferences. To further address this, we have built a phylogeny using IQ-TREE, as described previously, and partitioned the tree in 6 monophyletic clades (B_CS1 through B_CS6). These clades were examined for outlier sequences whose genetic divergence and sampling date were incongruent using TempEst software version 1.5.2 (**Rambaut et al, 2016**).

### Bayesian evolutionary inference of SARS-CoV-2 detected in Portugal

A total of 1275 SARS-CoV-2 genome sequences (obtained from positive samples collected until March 31^st^, 2020) from Portugal were analyzed in this study (INSA’s collection, as of July 23^rd^, 2020; Table S1). Our interest lies in estimating the viral evolutionary history and spatial diffusion process of the early epidemics in the country. Travel history data is of particular importance when analyzing low diversity data, such as that for SARS-CoV-2, using Bayesian joint inference of sequence and location traits because sharing the same location state can contribute to the phylogenetic clustering of taxa (**Lemey et al, 2020**). For each of the datasets (A, and B_CS1 through B_CS6), we performed a joint genealogical and phylogeographic inference of time-measured trees using Markov chain Monte Carlo (MCMC) sampling implemented in the Bayesian Evolutionary Analysis Sampling Trees (BEAST) package (**Suchard et al, 2018**). We applied a Hasegawa-Kishino-Yano 85 (HKY85) (**Hasegawa et al, 1985**) substitution model with gamma-distributed rate variation among sites (Shapiro et al, 2006). We used an uncorrelated lognormal relaxed molecular clock to account for evolutionary rate variation among lineages (**Drummond et al, 2006**) and specified an exponential growth coalescent prior in our analyses.

To integrate the travel history information obtained from (returning) travelers, we followed Lemey et al. (2020) (**Lemey et al, 2020**) and augmented the phylogeny with ancestral nodes that are associated with a location state (but not with a known sequence), and enforced the ancestral location at a point in the past of a lineage. We specified normal prior distributions on the travel times informed by an estimate of time of infection and truncated to be positive (back-in-time) relative to sampling date. Specifically, we use a period of 14 days (incubation period of 99% of patients (**Lauer et al, 2020**) where travel history information was collected for all recent movements, and a period between symptom onset and testing with an estimated mean of 4.70 days for the patients in the INSA cohort, and a standard deviation of 4.06 days to incorporate the uncertainty on the period between symptom onset and testing. The location traits associated with taxa and with the ancestral nodes were modeled using a bidirectional asymmetric discrete diffusion process (**Lemey et al, 2009**). We ran and combined at least eight independent MCMC analyses for 50 million generations, sampling every 50000th generation and removed 10% as chain burn-in. Stationarity and mixing was investigated using Tracer software version 1.7.1 (**Rambaut et al, 2018**), making sure that effective sample sizes for the continuous parameters were greater than 100. We used the high-performance computational capabilities of the Biowulf cluster at the National Institutes of Health (Bethesda, MD, USA) (http://biowulf.nih.gov) to perform these analyses. Portuguese clusters were assumed for phylogeographic summaries if their topology posterior probability was equal or higher than 0.001. If the excluded genomes had known travel history, they were re-integrated along with same-cluster sequences if those did not cluster in a clade-defining polytomy (recovered a total of 14 sequences, 7 of them with travel history).

### Real-time data sharing of SARS-CoV-2 genetic diversity and geotemporal spread in Portugal

A website (https://insaflu.insa.pt/covid19) was launched on March 28, 2020 for real-time data sharing on SARS-CoV-2 genetic diversity and geotemporal spread in Portugal. This site gives access to “situation reports of the study and provides interactive data navigation using both Nextstrain (https://nextstrain.org/) (**Hadfield et al, 2018**) and Microreact (https://microreact.org/) (**Árgimon et al, 2016**) tools. As of July 23^rd^, 2020, genomic and phylogenetic analysis were performed using the SARS-CoV-2 Nextstrain pipeline version from March 23, 2020 (https://github.com/nextstrain/ncov), with slight modifications (**Borges et al, 2020**). For data navigation, an IQ-TREE-derived (**Nguyen et al, 2015**) phylogenetic tree enrolling the 1275 studied sequences, and the associated metadata, can be visualized interactively at https://microreact.org/project/cM6KURnU7rUpqdAnBq5DAf/a2d3840e.

### Data availability

SARS-CoV-2 genome sequences generated in this study were uploaded to the GISAID database (https://www.gisaid.org/). Accession numbers can be found in Supplementary material (**Table S1**).

## RESULTS

### Epidemiological trends and circulating diversity during the early COVID-19 pandemic in Portugal

The first COVID-19 confirmed cases in Portugal were reported on March 2, 2020 after laboratory confirmation by the National Institute of Health (INSA) Doutor Ricardo Jorge. As the COVID-19 epidemic progressed in the country, INSA, as the National Reference Laboratory, gradually supported and validated the extension of the laboratory network throughout the country, where 37 laboratories were already set up to perform molecular testing of SARS-CoV-2 by March 31, 2020. With COVID-19 cases exponentially increasing in Portugal (**Figure 1A**) and Europe (with alarming trends in Italy and Spain) (**Dong et al, 2020**), Portugal adopted rigid public health measures, including suspension of flights from/to Italy (March 10, 2020), closure of land borders and schools (March 16, 2020), suspension of flights to non-EU countries and general lockdown (March 18, 2020). INSA rapidly implemented and coordinated the nationwide genomic surveillance of SARS-CoV-2 (https://insaflu.insa.pt/covid19) with a particular focus on providing a comprehensive picture of the introductions, genetic diversification and propagation of SARS-CoV-2 during the early-stage pandemic at a country scale. A total of 1275 SARS-CoV-2 positive samples collected in March were successfully subjected to virus genome sequencing (**Table S1**), which corresponds to 15.5% (1275/8251) of all COVID-19 cases confirmed during the first month of the pandemic in Portugal (**Figure 1B**). The viral genomic sequence sampling ranged from 10.7% (523/4910) in the Northern Health Administration region (the region with highest number of confirmed cases) to 85.4% (41/48) in Madeira Archipelago (the region with lowest number of confirmed cases) (**Figure 1C**). Despite the delayed epidemic trajectory in comparison with other European countries, Portugal was among the countries generating the highest volumes of SARS-CoV-2 genomic data during early COVID-19 pandemic (**Figure 1D; Table S2**).

**Figure 1.**
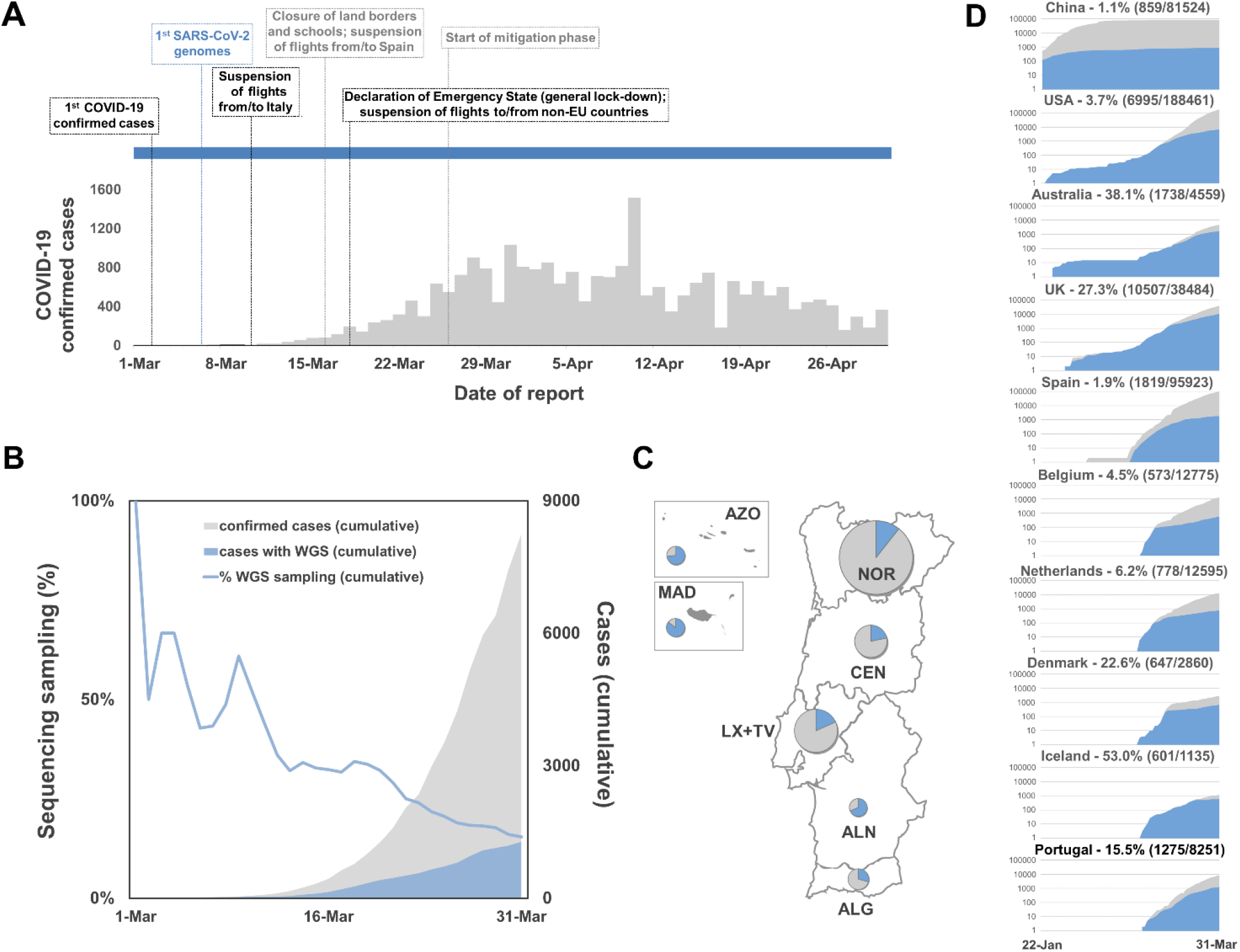
Overview of the COVID-19 confirmed cases and SARS-CoV-2 genome sequencing sampling in Portugal during the early phase of the pandemic. **A**. Daily reported COVID-19 confirmed cases in Portugal and key milestones during the early phase of the pandemic (source: Directorate-General of Health, https://covid19.min-saude.pt/relatorio-de-situacao/). **B**. Area plots (right y-axis) reflect the cumulative total number of COVID-19 confirmed cases (gray) and SARS-CoV-2 genome sequences (blue) reported/obtained in Portugal, until March 31, 2020. The blue line (right y-axis) displays the “sequencing sampling”, i.e. the proportion of confirmed cases with SARS-CoV-2 genome data during the same period. **C**. SARS-CoV-2 sequencing sampling generated by Health Administration region until March 31, 2020 (circles are proportional to the number of confirmed cases by Region, with the blue representing the proportion of samples with SARS-CoV-2 genome data). NOR, Northern region; CEN, Central region; LX+TV, Lisbon and Tagus Valley region; ALN, Alentejo; ALG, Algarve; AZO, Autonomous Region of Azores; MAD, AZO, Autonomous Region of Madeira. **D**. Area plots reflect the cumulative total number of COVID-19 confirmed cases (gray) and SARS-CoV-2 genome sequences (blue) reported by country between January 22 and March 31, 2020 for the top ten countries with the highest number of genomes with collection date until March 31, 2020 (available on GISAID by 8th August 2020). A log10 scale y-axis was used for visualization purposes. Countries are ordered according to the date of the first reported COVID-19 case.

The relative frequency trends of SARS-CoV-2 clades and lineages in Portugal during March 2020 (**Figure 2**) did not differ significantly from the scenario observed at the European level (**Alm et al, 2020**). Most of the analysed genomes (89.8%) belong to the 20A/G, 20B/GR and 20C/GH clades (Nextstrain/GISAID nomenclatures), which display, among other genetic markers, the D614G amino acid replacement in the Spike protein. The SARS-CoV-2 G614 variant was most likely introduced in Europe by mid-January / early February 2020 and became dominant at a worldwide level (**Nadeau et al, 2021**).

**Figure 2.**
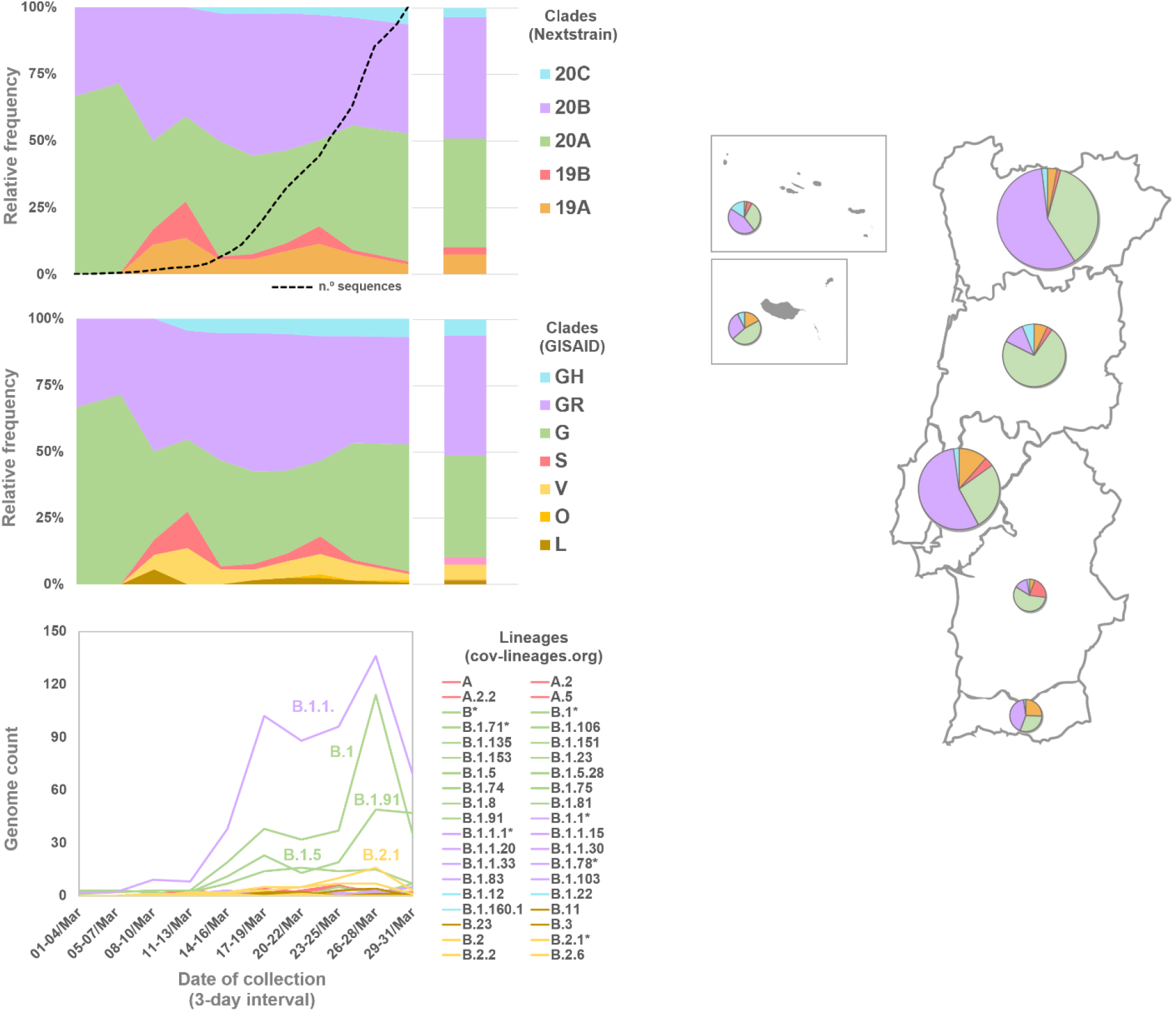
Frequency trends of SARS-CoV-2 clade and lineages in Portugal during March 2020 (n = 1275), according to the following nomenclature approaches: Nextstrain (https://clades.nextstrain.org/); GISAID (https://www.gisaid.org/), and Phylogenetic Assignment of Named Global Outbreak LINeages (cov-lineages.org) (https://pangolin.cog-uk.io/). Area plots reflect the Nextstrain/GISAID clade relative frequency by collection date (using 3-day interval with exception 1-4 March interval), while bar plots reflect the global relative frequency observed in March 2020. For the sake of graphical simplicity, the trajectory of the cov-lineages is shown as genome count instead of relative frequency. Pango lineages are colored according to the corresponding GISAID clades (when 100% congruence). For incongruent cases (labeled by an asterisk), the color reflects the most abundant GISAID clade within a given Pango lineage. The dashed line in the Nextstrain classification panel reflects the cumulative number of COVID-19 confirmed cases as shown in Figure 1. The map shows the relative frequency of Nextstrain clades by Health Administration region (circles are proportional to the number of confirmed cases by region) in March 2020. Classification is detailed in **Table S1**. The phylogeny, clade/lineage classification and geotemporal distribution can be visualized interactively at https://microreact.org/project/cM6KURnU7rUpqdAnBq5DAf/a2d3840e.

Clades 19A/L/V/O (dominant during early pandemic in China) and 19B/S (rare in Europe, with the notable exception of Spain) (**Alm et al, 2020**) were found at the relative frequencies around 7.5% and 2.8% in March, respectively, and showed a decreasing frequency trend similarly to what has been observed at the global level. When applying the Pango lineages (cov-lineages.org) nomenclature, the main B lineage (roughly covering 19A/20A/20B/20C Nextstrain clades) was dominant in March, showing ample diversification into sublineages. Among these, it is highlighted the increased frequency of the B.1.1 sublineages (roughly corresponding to clade 20B/GR), as well as of the B.1, B.1.5 and B.1.91 lineages (all mostly including 20A/G virus). Of note, the B.1.91 sub-lineage, and part of its ancestor sublineage B.1, correspond to the Spike G614+Y839 variant that massively disseminated in Portugal during the early epidemic (22% and 59% of the sampled genomes from the North and Center regions of Portugal by April 30^th^) (**Borges et al, 2020**).

### Introductions and early spread of SARS-CoV-2 in Portugal

In order to assess the origins and measure the number of SARS-CoV-2 introductions in Portugal prior March 31, 2020, we performed Bayesian phylodynamic analyses integrating the rich travel history data. Travel history could be collected for 619 (48.5%) out of 1275 confirmed cases detected in March, with 128 (10.0%) reporting international travels within the potential incubation period (i.e. 14 days before clinical onset date) and 491 (38.5%) reporting no travels within the same period (**Figure 3A**).

**Figure 3.**
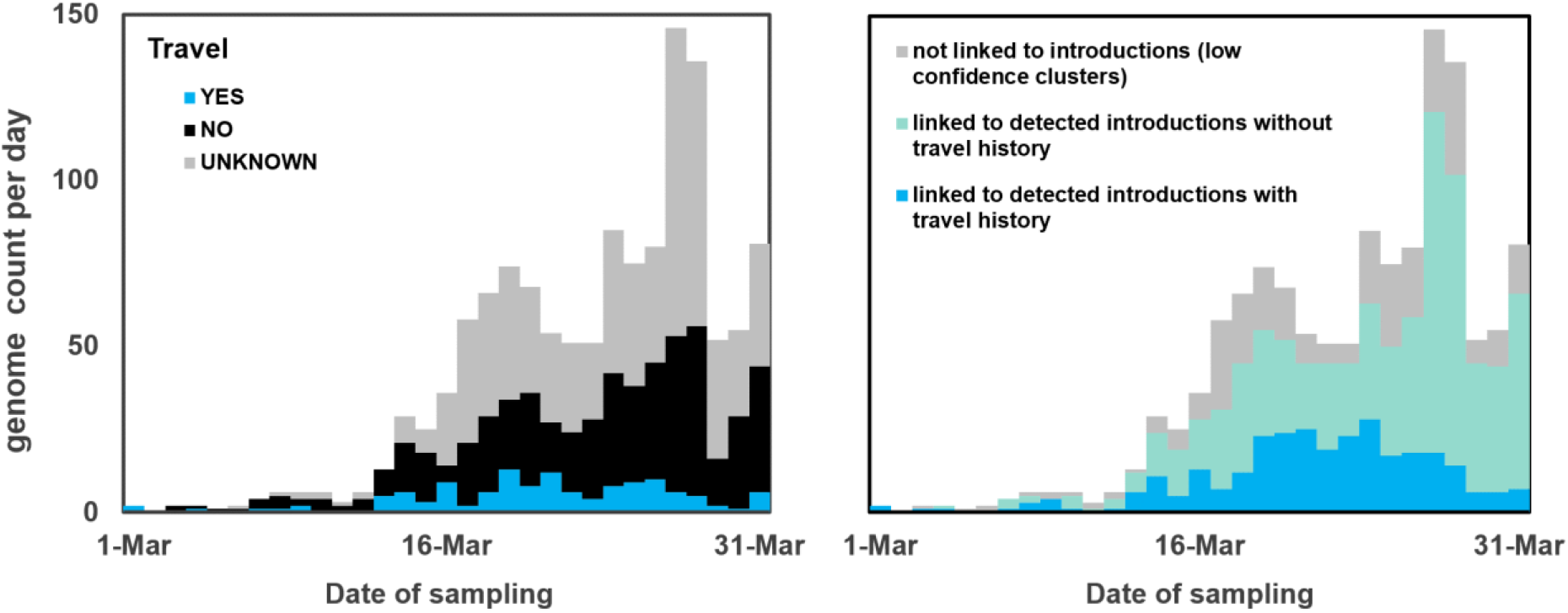
Overview of sequences linked to detected introductions, stratified by travel history data. **A**. Histogram of COVID-19 confirmed cases with SARS-CoV-2 genome sequencing data stratified by travel history. **B**. Histogram with sequence counts stratified by respective reconstruction linked with clades that include sequences from cases with (blue) or without (green) known travel history. Sequences belonging to BEAST clades with low topology posterior probability are shown in gray.

Overall, we detected 277 clades (16 from lineage A and 261 from lineage B) representing SARS-CoV-2 introductions in Portugal until March 31, 2020, involving a total of 979 out of the 1275 sequences analysed. Of these, 296/979 genomes belong to clades that include sequences from cases with known travel history (Figure 3B). Overall, the phylogeographic reconstruction revealed that UK, France and Spain seeded 69% of all introductions into Portugal (**Figure 4**), mostly to the Lisbon and Tagus Valley region, which was estimated to have received approximately 30% of all introductions, followed by the North (27%), Center (10%), Algarve (4 %), Azores (2%), Alentejo (2%) and Madeira (1%) regions (**Figure 5 and 6A; Table S4**).

**Figure 4.**
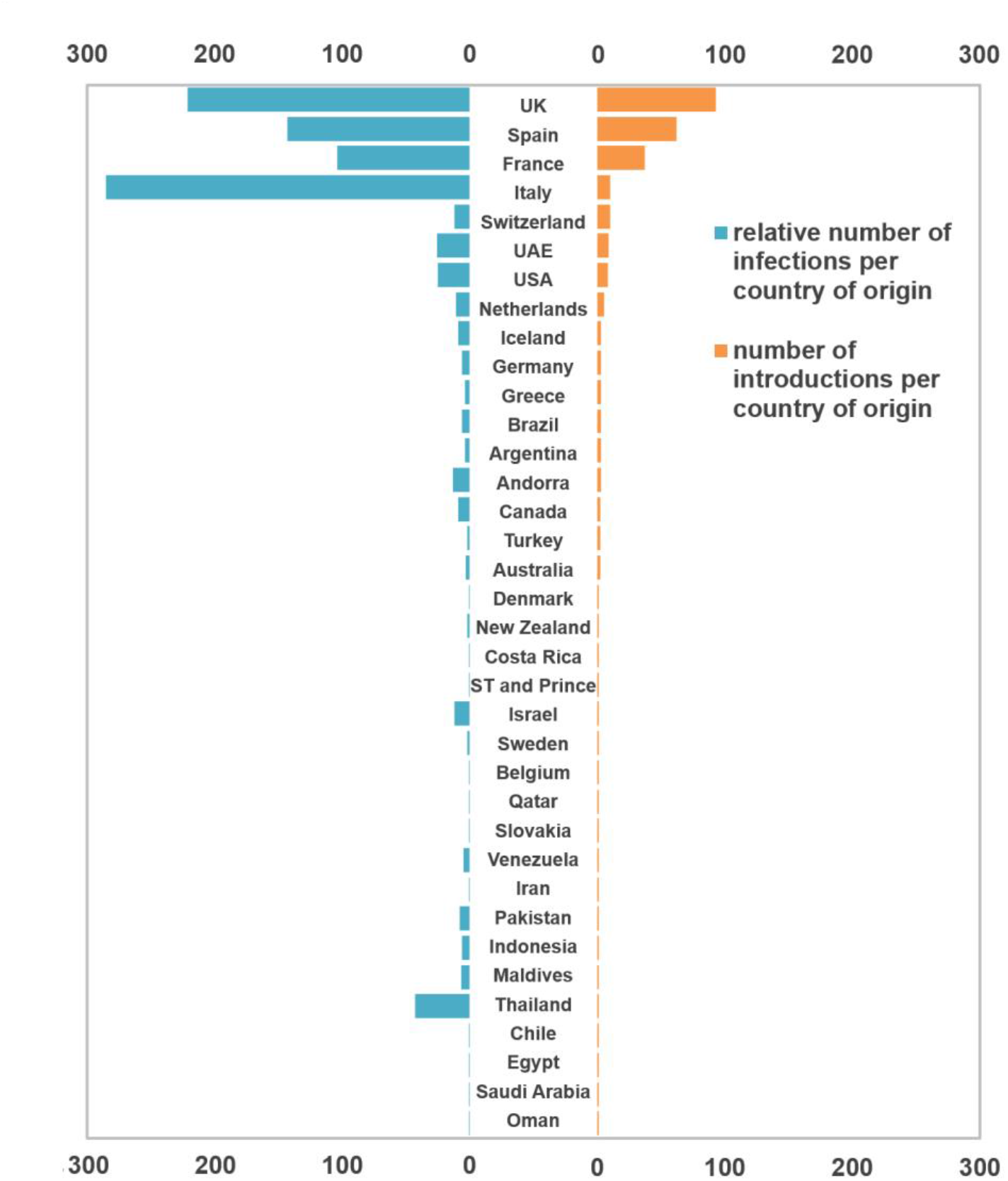
Number and size of SARS-CoV-2 introductions per country. Bar plots represent the number of introductions by country of origin (black) and respective total number of cases (blue).

**Figure 5.**
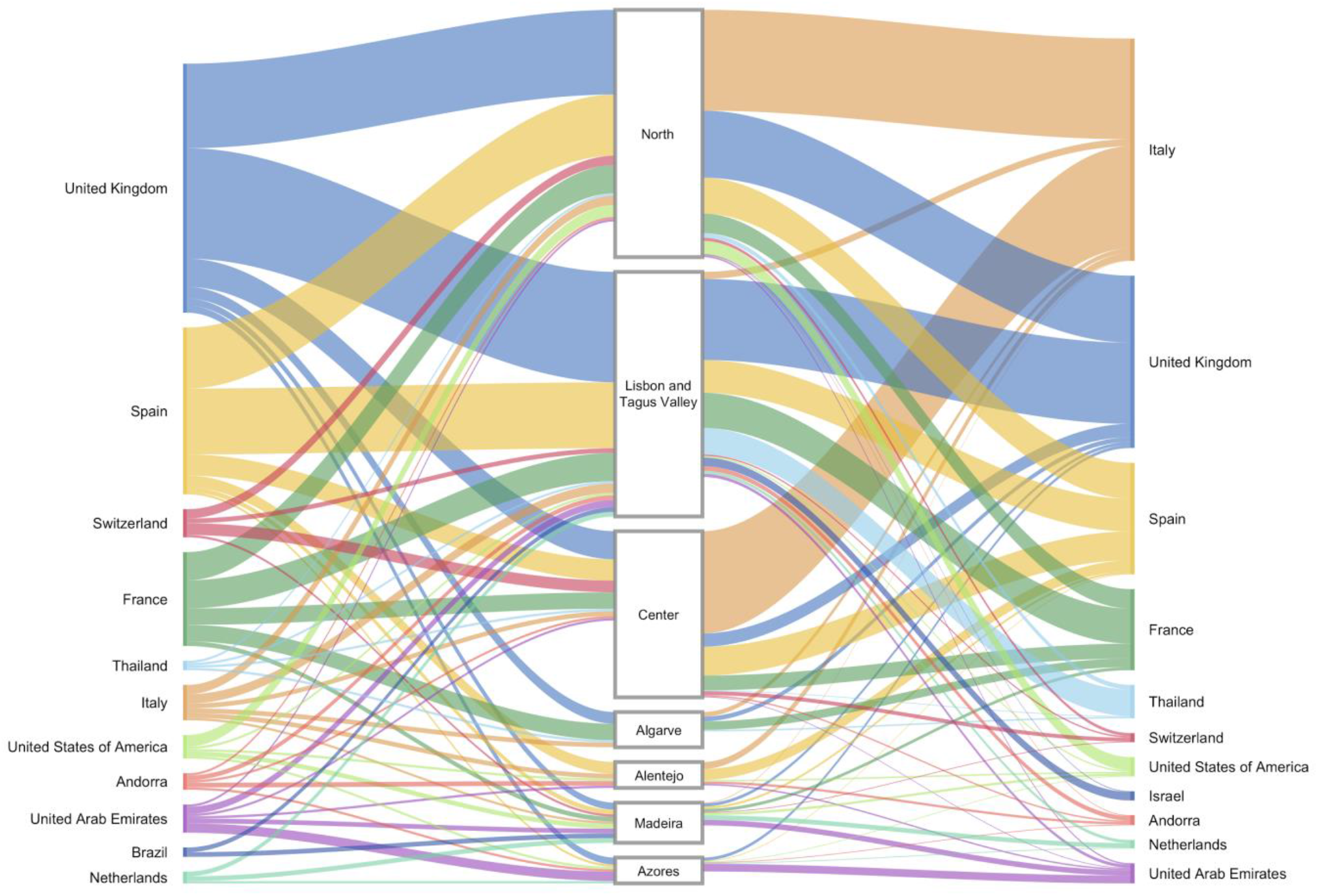
Sankey plots summarizing the viral flow into Portuguese Health Administration regions. **Left**. The plot shows the relative number of transitions between countries of origin and different regions in Portugal. For summaries that show all transitions to and from all connected locations, we refer to the **Table S4. Right**. The plot shows the relative number of infections generated by introductions from countries of origin and different regions in Portugal. For summaries that show the size of all introductions to and from all connected locations, we refer to the **Table S5**. For both panels, for graphics simplicity, we present the eleven countries linked with most introductions (left) and relative number of infections generated (right), estimated across phylogenies inferred for the whole dataset, i.e., including both Pango lineages A and B (thin – low proportion of number/size of viral introductions attributed to this source; thick – high proportion of number/size of viral introductions attributed to this source). We note that there is no temporal order for the transitions involved.

**Figure 6.**
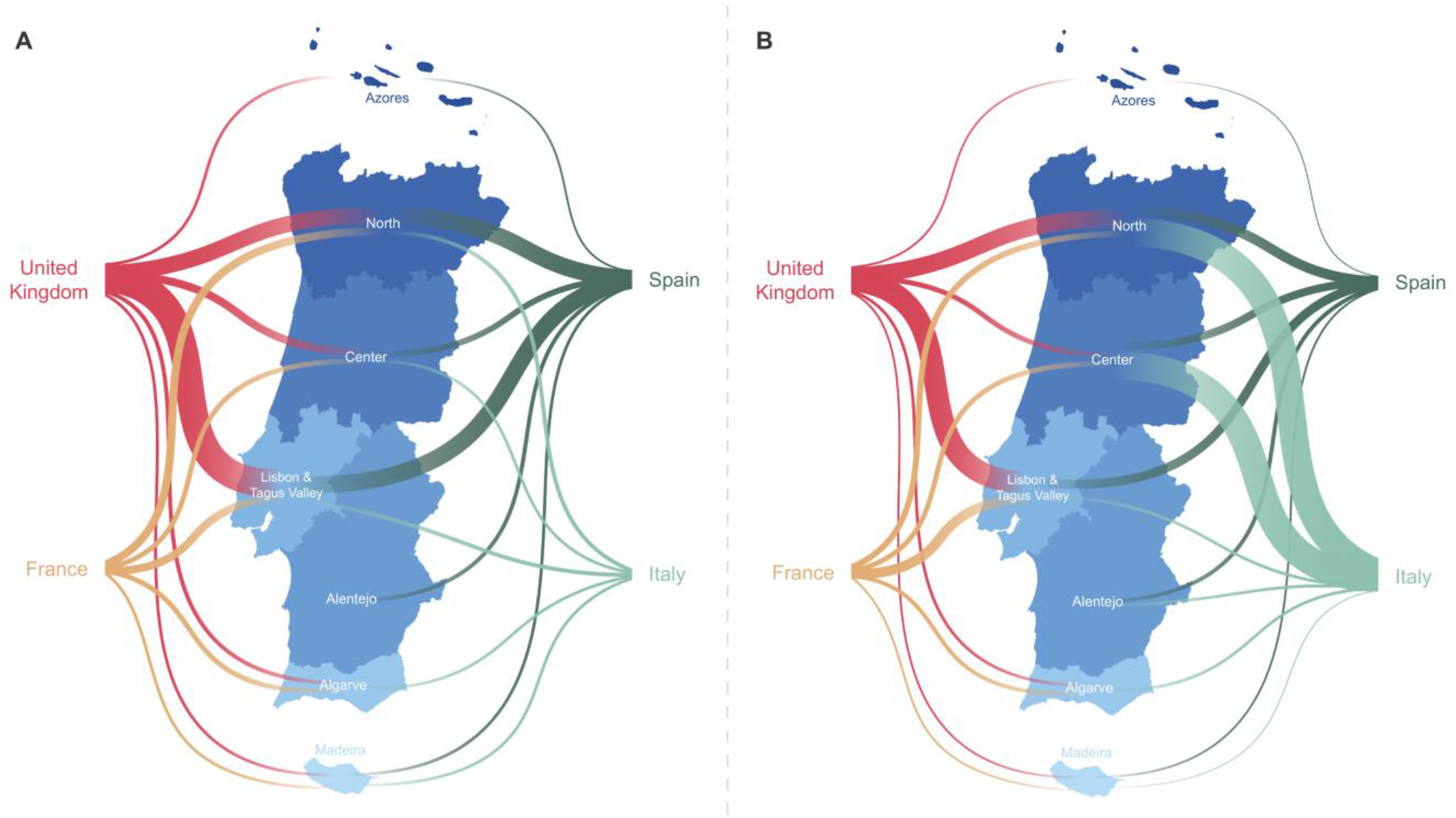
Map summarizes the viral flow into Portuguese Health Administration regions. **A**. The plot shows the relative number of transitions between countries of origin and different regions in Portugal. For summaries that show all transitions to and from all connected locations, we refer to the **Table S4. B**. The plot shows the relative number of infections generated by introductions from countries of origin and different regions in Portugal. For summaries that show the size of all introductions to and from all connected locations, we refer to the **Table S5**. For both panels, for graphics simplicity, we present the four countries linked with most introductions (left) and relative number of infections generated (right), estimated across phylogenies inferred for the whole dataset, i.e., including both Pango lineages A and B (thin – low proportion of number/size of viral introductions attributed to this source; thick – high proportion of number/size of viral introductions attributed to this source). We note that there is no temporal order for the transitions involved.

We also examined the number of infections generated by introductions from different countries (**Figure 5 and 6B; Table S5**). Interestingly, despite the UK, France and Spain having seeded the majority of introductions, at least one seeding event with travel history from Italy generated the largest outbreak recorded in early stages of the COVID-19 epidemic in Portugal (cfr. below) (**Borges et al, 2020**). We noticed a similar pattern for an introduction from Thailand, however this event had a low supported topology posterior probability (**Figure 5 and 6B; Table S5**).

The spatial analysis reconstructed 16 introductions of lineage A into Portugal encompassing 33 sequences from Portugal. The majority of the viral flow was seeded by Spain (69%), with 41% of all of these being introduced into Lisbon & Tejo Valley, 12% into the Center and Alentejo regions, respectively, and 6% into the North region. The remainder of all lineage A seeding events were estimated to have originated from Australia, France, Italy, Oman, Saudi Arabia and were mostly captured by the ability to integrate travel history data into the phylogeographic reconstruction. The introductions resulted in transmission clusters of varying size, with the largest, seeded by Spain, generating a transmission chain of at least 7 cases in Alentejo.

A similar phylogeographic analysis for lineage B resulted in the reconstruction of 261 introductions covering 946 Portuguese genomes. The majority of lineage B introductions were seeded by the UK (36%), Spain (20%) and France (14%). The introductions resulted in transmission clusters of varying size. The largest was seeded by Italy and generated a transmission chain of at least 252 cases in the North and Center regions. Switzerland (4%), UAE (3%), USA (3%), Netherlands (2%) also contributed to the remainder of lineage B introductions and establishment in the country.

Comparison of the times to the most recent common ancestor (tMRCA) across BEAST clades representing seeding events into Portugal and the date of collection of the most recent sampled genome in that clade revealed a median time lag between tMRCA (representing the earliest an introduction could have occurred) and genomic surveillance of 20 days (range = 1 and 68 days; **Figure 7**). In particular, the tMRCA of most clades (∼44%) occurred between the last week of February and the first week of March, suggesting most introductions were seeded by SARS-CoV-2 genetic variants that were circulating at the same time in the estimated country of origin. The temporal reconstruction estimated the earliest introduction of lineage A into Portugal to have a time to the most recent common ancestor (tMRCA) as early as February 12, 2020, from Spain to the Lisbon & Tejo Valley region. The earliest seeding event for lineage B has a tMRCA on January 21, 2020 via Spain into the North region. Importantly, most clades with estimated tMRCA before February belong to Nextstrain 20B clade. Because of both the high abundance of 20B sequences in the dataset and the low genetic diversity within this clade at the time (leading to polytomy-rich topologies), the tMRCAs of these introductions might represent the tMRCA of 20B clade per se, rather than the importation date of the variant into Portugal. Notwithstanding, our data suggests a period of up to six weeks before the first COVID-19 confirmed cases, where undetected transmission likely occurred (**Figure 7**).

**Figure 7.**
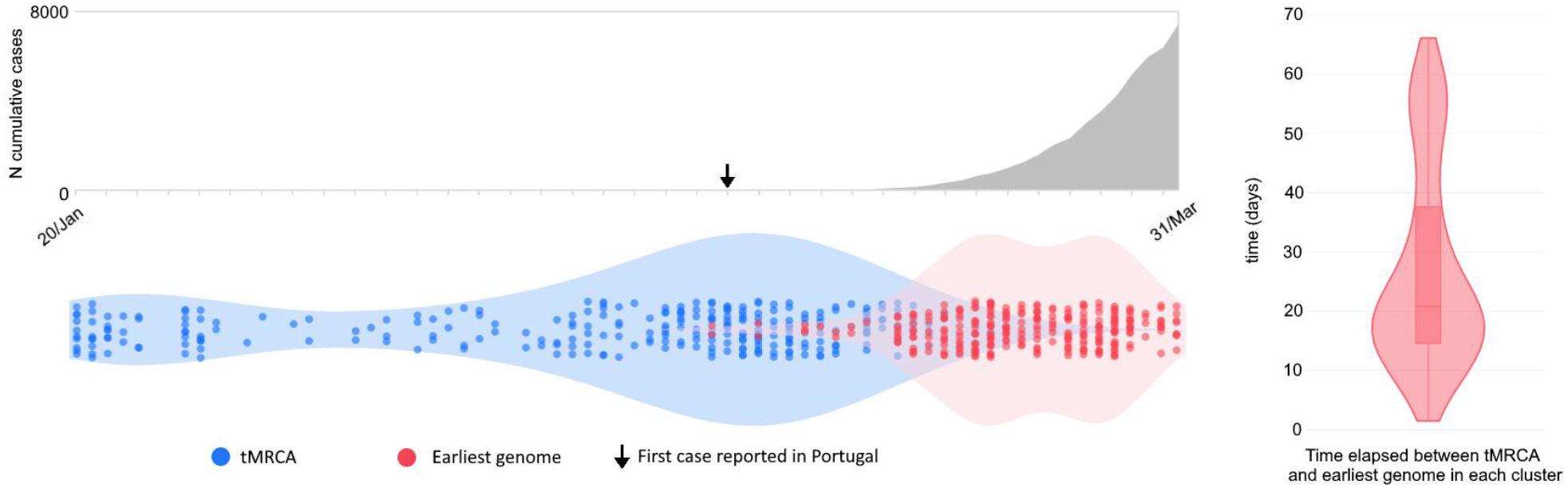
Cryptic transmission of SARS-CoV-2 in Portugal revealed by genomic epidemiology. **A**. Number of cumulative cases over time. **B**. Violin plots represent the date of the earliest genome in a Portuguese clade (red) and the time to the most recent common ancestor (tMRCA in blue). **C**. Violin plot and point density depict the time lag between the introduction and the first surveilled genome.

## DISCUSSION

The present study provides a comprehensive description of the early establishment of the COVID-19 pandemic in Portugal. We detected multiple independent SARS-CoV-2 introductions, mostly from European countries (namely the UK, Spain, France, Italy and Switzerland), which was broadly consistent with the available travel history data, as well as with the countries with most frequent flight connectivity (**INE, 2020**) and/or with the highest number of Portuguese immigrants (**UN, 2019**).

The genomic surveillance efforts culminated into the generation of 1275 SARS-CoV-2 whole genomes, that represent 15.5% of all confirmed cases in March, making Portugal the country generating the 5th highest number of SARS-CoV-2 genomic sequences during the early COVID-19 pandemic. In particular, our data uncovered the importation of extensive SARS-CoV-2 genomic diversity during the early epidemic in Portugal, as supported by the identification of at least 277 seeding events, covering all Nextstrain/GISAID clades and dozens of Pango lineages. Despite the vast genetic diversity introduced, 32% of the 1275 analysed sequences could be linked to just ten out of the 277 detected introductions. In particular, a single introduction (from Italy) of a Spike Y839 variant represented about 20% of all sampled genomes collected until the end of March (**Borges et al, 2020**). Moreover, 56% (155/277) of BEAST clades involve one single sequence (singletons), thus suggesting that most introductions have not seeded substantial local transmission.

In general, these data suggest that the implemented preventive and early control measures have been successful in mitigating the establishment of community transmission from most SARS-CoV-2 independent introductions. On the other hand, it also highlights that the few introductions not captured by public health surveillance and control can still seed large community transmission events and give rise to a significant number of cases. This underlies the challenges in defining a public health strategy aimed at preventing sustained community transmission while maintaining open borders and global connectivity. More stringent border restrictions, including flight and land borders closure, were implemented after March 10, when 87% (240/277) of the detected introductions were estimated to have already occurred. Portugal presents a geopolitical and demographic context involving high circulation of both migrant workers and tourists, and close international networks with countries with multiple countries from different latitudes (especially from Europe, South America and Africa). In light of the current knowledge of the COVID-19 pandemic dynamics and the challenging trade-off between the extension of restriction measures and the countries’ capacity to control the virus dissemination, it is now clear that restrictions in circulation should have been implemented before the confirmation of first COVID-19 cases in Portugal. In fact, despite the delay on the epidemic start in Portugal in comparison with other European countries, which certainly contributed to a more favorable situation during the pandemic’s first epidemic wave, the implementation of measures (close borders, travelers testing, etc.) in an earlier time frame could have largely minimized the number of introductions and subsequent virus expansion.

The lag in detection observed between the estimated time of the most recent common ancestor and the collection date of the earliest sample of each BEAST clade denotes that cryptic transmission might have occurred to some extent. Nonetheless, some of the earlier estimated tMRCAs (especially those in January) were associated with clades with low topological support and thus should be interpreted with caution. Although most introductions were estimated to have occurred during the last week of February and the first week of March 2020, SARS-CoV-2 was silently circulating in Portugal several weeks before the first confirmed local cases on March 2, 2020. For instance, 12 out of 19 sequences collected during the first week of the epidemic were obtained from patients with no travel history, including patients linked to the introduction leading to the highest number of cases during the first wave.

Phylodynamic modeling relies on the accumulation of genetic diversity over time for estimation of the evolutionary rate and timing of other relevant events. Being a recently emerged pathogen with a lower substitution rate (in comparison with other RNA viruses) and a large genome, it is challenging to study the early dynamics of SARS-CoV-2, as a large proportion of sequences collected across time and space are very closely related genetically, thus complicating the reconstruction of the phylogenetic topology. To address this, we opted to focus on a subset of the results with higher topological support. Furthermore, due to the computational burden required for phylodynamic analysis, it is unfeasible to include all available genomic sequences. One limitation of this and other studies trying to infer the origin of introductions is the sequencing sampling bias by country. For this reason, we resorted to filtering and subsampling strategies that aim at reducing the number of genomes that do not significantly contribute to the evolutionary or phylogeographic reconstructions. Despite being less pronounced, there were still discrepancies in the number of genomes included across countries. For instance, it is very likely that the number of introductions via the UK (the country generating the highest volume of sequences) is overestimated, while the number of introductions from countries with none or few available genome sequences is underestimated. However, the integration of travel history during the phylodynamic inferences allows for a more accurate reconstruction of the spatial pathways from both over and undersampled locations. Additionally, using this recently developed model provides insight into the genetic diversity circulating in countries for which genomic surveillance is still lacking. This makes the present work among the first to apply this novel model, which allowed us to gain insight into the early circulating diversity in countries like Saint Thomas and Prince, for which (as of February 15, 2021) there are no sequences available, and other undersampled countries such as the United Arabe Emirates and Qatar.

Overall, our findings highlight the use of genomic data to trace the introduction and spread of an emerging virus, showing the need of systematic, continuous and geographically representative genomic surveillance to detect and monitor the emergence and dissemination of biologically and/or epidemiologically relevant variants. Together with open data sharing, the timely generation of SARS-CoV-2 genomic data has been shown to be an invaluable tool to guide national and international public health authorities towards the identification and control of highly transmissible and/or immune evading variants (**ECDC, 2021**).

In this context, by laying the foundation of the genomic epidemiology of SARS-CoV-2 in Portugal, involving an unprecedented collaborative effort at national and international levels, this work, and subsequent capacity building, was pivotal for Portugal to respond to the current and upcoming needs of a genomic-informed surveillance and epidemiology of COVID-19, as strongly recommended by ECDC and WHO (**WHO, 2021; ECDC, 2021**).

## Supplementary Material

**Table S1. List of SARS-CoV-2 genome sequences (and respective Clade/lineage classification) from Portugal used in this study**. (Excel file)

**Table S2. Number of cumulative cases of COVID-19 by 31 March 2020, by country, and respective number of available SARS-CoV-2 genomes with collection date until 31 March (and available on GISAID as of 8 August 2020)**. Countries are shown by descending order of number of genomes and only countries with at least 30 COVID-19 reported cases and more than 2 genomes available are represented. Countries highlighted in bold are represented in **Figure 1D**. (Excel file)

**Table S3. GISAID acknowledgment table for the background dataset**. We acknowledge the original and submitting laboratories that generated the full-length viral genome sequences downloaded from GISAID (https://www.gisaid.org/) on August 6, 2020 with collection dates before April 1, 2020. (PDF file)

**Table S4. Heatmap with relative number of transitions between countries of origin and different regions in Portugal**. (Excel file)

**Table S5. Heatmap with relative number of infections generated by introductions from countries of origin and different regions in Portugal**. (Excel file)

**List of authors of the consortium entitled “Portuguese network for SARS-CoV-2 genomics”** (PDF file)

### Ethical Approval

This study was approved by the Ethical Committee (“Comissão de Ética para a Saúde”) of the Portuguese National Institute of Health. Designations of all genome sequences are fully anonymized, and no identifying information of the associated patients is provided.

## Supporting information

Portuguese network for SARS-CoV-2 genomics (Consortium)

## Data Availability

SARS-CoV-2 genome sequences generated in this study were uploaded to the GISAID database (https://www.gisaid.org/).

## Acknowledgments

We gratefully acknowledge to Sara Hill and Nuno Faria (University of Oxford) and Joshua Quick and Nick Loman (University of Birmingham) for kindly providing us with the initial sets of Artic Network primers for NGS; Rafael Mamede (MRamirez team, IMM, Lisbon) for developing and sharing a bioinformatics script for sequence curation (https://github.com/rfm-targa/BioinfUtils); Philippe Lemey (KU Leuven) for providing guidance on the implementation of the phylodynamic models; Joshua L. Cherry (National Center for Biotechnology Information, National Library of Medicine, National Institutes of Health) for providing guidance with the subsampling strategies; and all authors, originating and submitting laboratories who have contributed genome data on GISAID (https://www.gisaid.org/) on which part of this research is based. The opinions expressed in this article are those of the authors and do not reflect the view of the National Institutes of Health, the Department of Health and Human Services, or the United States government.

