## Supplementary material for "The early dynamics of the SARS-CoV-2 epidemic in Portugal": Portuguese network for SARS-CoV-2 genomics (Consortium)

### Supplementary Information

#### Authors list within the consortium entitled "Portuguese network for SARS-CoV-2 genomics"

All members (listed below) of the consortium entitled "Portuguese network for SARS-CoV-2 genomics" are authors of this manuscript.


| Author | Affiliation | Contact |
|---|---|---|
| Carlos Sousa | Laboratório De Biologia Molecular da Unilabs Portugal | |
| Pedro Cardoso | Laboratório De Biologia Molecular da Unilabs Portugal | |
| Carlos Cardoso | Laboratório de Análises Clínicas Dr Joaquim Chaves | |
| Laura Brum | Synlab Lisboa | |
| Lurdes Monteiro | Secção de Patologia Molecular, Synlab Lisboa | |
| Cristina Toscano | Laboratório de Microbiologia e Biologia Molecular do Centro Hospitalar de Lisboa Ocidental | |
| Maria Ana Pessanha | Laboratório de Microbiologia e Biologia Molecular do Centro Hospitalar de Lisboa Ocidental | |
| Ana Paula Dias | Laboratório de Microbiologia e Biologia Molecular do Centro Hospitalar de Lisboa Ocidental | |
| João Dias | Laboratório de Microbiologia e Biologia Molecular do Centro Hospitalar de Lisboa Ocidental | |
| Maria Helena Ramos | Serviço de Microbiologia, Centro Hospitalar do Porto | |
| Ana Constança | Serviço de Microbiologia, Centro Hospitalar do Porto | |
| Agostinho José S. Lira | Centro Hospitalar de Vila Nova de Gaia e Espinho | |
| Filomena Lacerda | Centro Hospitalar de Vila Nova de Gaia e Espinho | |
| Luis Marques Silva | Centro Hospitalar de Vila Nova de Gaia e Espinho | |
| Maria Matos Figueiredo | Centro Hospitalar de Vila Nova de Gaia e Espinho | |
| Nair Seixas | Centro Hospitalar de Vila Nova de Gaia e Espinho | |
| Jorge Meneses | Centro Hospitalar de Vila Nova de Gaia e Espinho | |
| Paulo Leandro | Centro Hospitalar de Vila Nova de Gaia e Espinho | |
| Alexandra Estrada | Serviço de Patologia Clínica, Hospital de Braga | |
| Fernando Branca | Serviço de Patologia Clínica, Hospital de Braga | |
| Aida M. Sousa Fernandes | Laboratório Regional de Saúde Pública Drª Laura Ayres - ARS Algarve | |
| Paula Barbeiro | Serviço de Patologia Clinica - Unidade Local de Saúde Litoral Alentejano | |
| Inna Slobidnyk | Serviço de Patologia Clinica - Unidade Local de Saúde Litoral Alentejano | |
| Maria João Peres | Laboratório de Imunologia e Biologia Molecular, Centro Hospitalar de Setúbal | |
| Rita Côrte-Real | Laboratório Biologia Molecular- Serviço de Patologia Clínica,Centro Hospitalar Universitário Lisboa Central | |
| Madalena Almeida Santos | Laboratório Biologia Molecular- Serviço de Patologia Clínica,Centro Hospitalar Universitário Lisboa Central | |
| Olga Costa | Laboratório Biologia Molecular- Serviço de Patologia Clínica,Centro Hospitalar Universitário Lisboa Central | |
| Conceição Godinho | Laboratório Biologia Molecular- Serviço de Patologia Clínica,Centro Hospitalar Universitário Lisboa Central | |
| Paula Branquinho | Laboratório Biologia Molecular- Serviço de Patologia Clínica,Centro Hospitalar Universitário Lisboa Central | |

| | | |
|---|---|---|
| Lurdes Lopes | Laboratório Biologia Molecular- Serviço de Patologia Clínica,Centro Hospitalar Universitário Lisboa Central | |
| Paula Soares | Laboratório Biologia Molecular- Serviço de Patologia Clínica,Centro Hospitalar Universitário Lisboa Central | |
| Lidia Santos | Laboratório Biologia Molecular- Serviço de Patologia Clínica,Centro Hospitalar Universitário Lisboa Central | |
| Patricia Miguel | Laboratório Biologia Molecular- Serviço de Patologia Clínica,Centro Hospitalar Universitário Lisboa Central | |
| Isabel Dias | Laboratório Biologia Molecular- Serviço de Patologia Clínica,Centro Hospitalar Universitário Lisboa Central | |
| Isabel Fernandes | Laboratório Biologia Molecular- Serviço de Patologia Clínica,Centro Hospitalar Universitário Lisboa Central | |
| Sónia Rodrigues | Laboratório Biologia Molecular- Serviço de Patologia Clínica,Centro Hospitalar Universitário Lisboa Central | |
| Fátima Vale | Serviço de Patologia Clínica, Unidade Local de Saúde da Guarda | |
| Joana Ramos | Serviço de Patologia Clínica, Unidade Local de Saúde da Guarda | |
| Rita Gralha | Serviço de Patologia Clínica, Unidade Local de Saúde da Guarda | |
| Patricia Fonseca | Serviço de Patologia Clínica, Unidade Local de Saúde da Guarda | |
| Nelson Ventura | Serviço de Patologia Clínica, Unidade Local de Saúde da Guarda | |
| Filomena Caldeira | Hospital Espírito Santo, Évora | |
| Margarida Farinha | Serviço de Patologia Clínica, Centro Hospitalar Tondela-Viseu | |
| Ana Caldas | Serviço de Patologia Clínica, Centro Hospitalar Tondela-Viseu | |
| Carina de Fátima Rodrigues | Centro de Investigação de Montanha, Instituto Politécnico de Bragança | |
| Maria Alice Pinto | Centro de Investigação de Montanha, Instituto Politécnico de Bragança | |
| António Albuquerque | Unidade Local de Saúde de Matosinhos | |
| Valquíria Alves | Unidade Local de Saúde de Matosinhos | |
| João Carlos Sousa | Life and Health Sciences Research Institute, School of Medicine, University of Minho, Braga | |
| Maria Isabel Veiga | Life and Health Sciences Research Institute, School of Medicine, University of Minho, Braga | |
| Diana Patrícia Pinto da Silva | Centro Médico da Praça | |
| Ricardo Filipe Romão Ferreira | Centro Médico da Praça | |
| Maria Beatriz Tomaz | Beatriz Godinho Saúde | |
| Alfredo Rodrigues | Beatriz Godinho Saúde | |
| Jácome Bruges Armas | Serviço Especializado de Epidemiologia e Biologia Molecular, Hospital de Santo Espírito, Ilha Terceira, Açores | |
| Ana Rita Couto | Serviço Especializado de Epidemiologia e Biologia Molecular, Hospital de Santo Espírito, Ilha Terceira, Açores | |
| Paula Valente | Departamento de Saúde Pública e Planeamento, ARS Alentejo | |
| Cláudia Nunes dos Santos | Centro de Estudos de Doenças Crónicas, Faculdade de Ciências Médicas, Universidade Nova de Lisboa | |
| Paulo Pereira | Centro de Estudos de Doenças Crónicas, Faculdade de Ciências Médicas, Universidade Nova de Lisboa | |
| José Alves | Serviço de Patologia Clínica - Hospital Dr. Nélio Mendonça - SESARAM | |
| Graça Andrade | Serviço de Patologia Clínica - Hospital Dr. Nélio Mendonça - SESARAM | |
| Ludivina Freitas | Serviço de Patologia Clínica - Hospital Dr. Nélio Mendonça - SESARAM | |
| Bruna R. Gouveia | Interactive Technologies Institute - LARSyS | |

| | | |
|---|---|---|
| Pedro Ramos | Secretaria Regional de Saúde e Proteção Civil - Governo Regional da Madeira | |
| Herberto Jesus | Instituto de Administração da Saúde da Madeira | |
| Maurício Melim | Instituto de Administração da Saúde da Madeira | |
| Hugo Sousa | Serviço de Virologia, Instituto Português de Oncologia do Porto | |
| Inês Baldaque | Serviço de Virologia, Instituto Português de Oncologia do Porto | |
| Daniela Silva | ALS Controlvet , Tondela | |
| Inês Gomes | ALS Controlvet, Tondela | |
| Eliana Costa | Serviço de Patologia Clínica, Centro Hospitalar de Trás-os-Montes e Alto Douro | |
| Sara Sousa | Serviço de Patologia Clínica, Centro Hospitalar de Trás-os-Montes e Alto Douro | |
| Ana Miguel Matos | Laboratório de Análises Clínicas da Universidade de Coimbra | |
| Miguel Babarro Jorreto | Serviço de Imunohemoterapia, Unidade Local de Saúde do Alto Minho | |
| Maria da Graça Maciel de Soveral | Serviço de Imunohemoterapia, Unidade Local de Saúde do Alto Minho | |
| Luís Silva | Serviço de Patologia Clínica, Hospital de Vila Franca de Xira | |
| Helena Ribeiro | Serviço de Patologia Clínica, Hospital de Vila Franca de Xira | |
| Rita Rodrigues | Serviço de Patologia Clínica, Hospital de Vila Franca de Xira | |
| Teresa Salvado | Serviço de Patologia Clínica, Hospital de Vila Franca de Xira | |
| Luisa Mota-Vieira | Unidade de Genética e Patologia Moleculares, Hospital do Divino Espirito Santo de Ponta Delgada, Ilha de S. Miguel, Açores | |
| Rita C. Veloso | Unidade de Genética e Patologia Moleculares, Hospital do Divino Espirito Santo de Ponta Delgada, Ilha de S. Miguel, Açores | |
| Claudia C. Branco | Unidade de Genética e Patologia Moleculares, Hospital do Divino Espirito Santo de Ponta Delgada, Ilha de S. Miguel, Açores | |
| Sónia Marta Santos Magalhães | Hospital Agostinho Ribeiro-Felgueiras | |
| Helena Rodrigues | Laboratório Dra Helena Rodrigues, Valença | |
| Francisca Rocha | Laboratório Dra Helena Rodrigues, Valença | |
| Sandra Paulo | Serviço de Patologia Clínica da Unidade Local de Saúde de Castelo Branco | |
| Mariana Martins | Serviço de Patologia Clínica da Unidade Local de Saúde de Castelo Branco | |
| Ricardo Rodrigues | Serviço de Patologia Clínica da Unidade Local de Saúde de Castelo Branco | |
| Mariana Viana | Centro Hospitalar Tâmega e Sousa, Penafiel | |
| Maria Calle Vellés | Centro Hospitalar Tâmega e Sousa, Penafiel | |
| Miguel Pinheiro | iBiMED/Universidade de Aveiro | |
| Miguel Fevereiro | Instituto Nacional de Investigação Agrária e Veterinária | |
| Ana Margarida Henriques | Instituto Nacional de Investigação Agrária e Veterinária | |
| Tiago Luís | Instituto Nacional de Investigação Agrária e Veterinária | |
| Cathy Paulino | Instituto Gulbenkian de Ciência, Oeiras | |
| João Costa | Instituto Gulbenkian de Ciência, Oeiras | |
| João Sobral | Instituto Gulbenkian de Ciência, Oeiras | |
| Susana Ladeiro | Instituto Gulbenkian de Ciência, Oeiras | |
| Jorge Machado | Instituto Nacional de Saúde Dr Ricardo Jorge (INSA), Lisboa | |
| Paula Bajanca-Lavado | Instituto Nacional de Saúde Dr Ricardo Jorge (INSA), Lisboa | |
| Maria José Borrego | Instituto Nacional de Saúde Dr Ricardo Jorge (INSA), Lisboa | |
| Líbia Zé-Zé | Instituto Nacional de Saúde Dr Ricardo Jorge (INSA), Lisboa | |

| | | |
|---|---|---|
| Nuno Verdasca | Instituto Nacional de Saúde Dr Ricardo Jorge (INSA), Lisboa | |
| Sílvia Lopo | Instituto Nacional de Saúde Dr Ricardo Jorge (INSA), Lisboa | |
| Rita de Sousa | Instituto Nacional de Saúde Dr Ricardo Jorge (INSA), Lisboa | |
| Maria João Gargate | Instituto Nacional de Saúde Dr Ricardo Jorge (INSA), Lisboa | |
| Susana Martins | Instituto Nacional de Saúde Dr Ricardo Jorge (INSA), Lisboa | |
| Isabel Lopes de Carvalho | Instituto Nacional de Saúde Dr Ricardo Jorge (INSA), Lisboa | |
| Célia Rodrigues Bettencourt | Instituto Nacional de Saúde Dr Ricardo Jorge (INSA), Lisboa | |
| Carla Roque | Instituto Nacional de Saúde Dr Ricardo Jorge (INSA), Lisboa | |
| Leonor Silveira | Instituto Nacional de Saúde Dr Ricardo Jorge (INSA), Lisboa | |
| João Rodrigues | Instituto Nacional de Saúde Dr Ricardo Jorge (INSA), Lisboa | |
| Ivone Água-Doce | Instituto Nacional de Saúde Dr Ricardo Jorge (INSA), Lisboa | |
| Rita Cordeiro | Instituto Nacional de Saúde Dr Ricardo Jorge (INSA), Lisboa | |
| Ana Pelerito | Instituto Nacional de Saúde Dr Ricardo Jorge (INSA), Lisboa | |
| Cristina Correia | Instituto Nacional de Saúde Dr Ricardo Jorge (INSA), Lisboa | |
| Vera Manageiro | Instituto Nacional de Saúde Dr Ricardo Jorge (INSA), Lisboa | |
| Raquel Rocha | Instituto Nacional de Saúde Dr Ricardo Jorge (INSA), Lisboa | |
| Raquel Neves | Instituto Nacional de Saúde Dr Ricardo Jorge (INSA), Lisboa | |
| Paula Palminha | Instituto Nacional de Saúde Dr Ricardo Jorge (INSA), Lisboa | |
| Cristina Veríssimo | Instituto Nacional de Saúde Dr Ricardo Jorge (INSA), Lisboa | |
| Elizabeth Pádua | Instituto Nacional de Saúde Dr Ricardo Jorge (INSA), Lisboa | |
| Rita Matos | Instituto Nacional de Saúde Dr Ricardo Jorge (INSA), Lisboa | |
| Susana Silva | Instituto Nacional de Saúde Dr Ricardo Jorge (INSA), Lisboa | |
| Alexandra Nunes | Instituto Nacional de Saúde Dr Ricardo Jorge (INSA), Lisboa | |
| Pedro Pechirra | Instituto Nacional de Saúde Dr Ricardo Jorge (INSA), Lisboa | |
| Inês Costa | Instituto Nacional de Saúde Dr Ricardo Jorge (INSA), Lisboa | |
| Mónica Oleastro | Instituto Nacional de Saúde Dr Ricardo Jorge (INSA), Lisboa | |
| Carla Feliciano | Instituto Nacional de Saúde Dr Ricardo Jorge (INSA), Lisboa | |
| Isabel Albergaria | Instituto Nacional de Saúde Dr Ricardo Jorge (INSA), Lisboa | |
| Fernanda Vilarinho | Instituto Nacional de Saúde Dr Ricardo Jorge (INSA), Lisboa | |
| Márcia Faria | Instituto Nacional de Saúde Dr Ricardo Jorge (INSA), Lisboa | |
| Margarida Vaz | Instituto Nacional de Saúde Dr Ricardo Jorge (INSA), Lisboa | |
| Patrícia Barros | Instituto Nacional de Saúde Dr Ricardo Jorge (INSA), Lisboa | |
| Raquel Rodrigues | Instituto Nacional de Saúde Dr Ricardo Jorge (INSA), Lisboa | |
| José Vicente Constantino | Instituto Nacional de Saúde Dr Ricardo Jorge (INSA), Lisboa | |
| Rita Macedo | Instituto Nacional de Saúde Dr Ricardo Jorge (INSA), Lisboa | |
| Raquel Sabino | Instituto Nacional de Saúde Dr Ricardo Jorge (INSA), Lisboa | |
| Idalina Ferreira | Instituto Nacional de Saúde Dr Ricardo Jorge (INSA), Lisboa | |
| Sónia Silva | Instituto Nacional de Saúde Dr Ricardo Jorge (INSA), Lisboa | |
| Anabela Vilares | Instituto Nacional de Saúde Dr Ricardo Jorge (INSA), Lisboa | |
